# Development and validation of electronic health record-based ascertainment of obsessive-compulsive disorder cases and controls

**DOI:** 10.1101/2025.08.05.25332874

**Authors:** Bo Wang, Tyne W. Miller-Fleming, Dongmei Yu, Donald Hucks, Emily Gantz, Rebecca Johnston, Angela Maxwell-Horn, Nancy Cox, James Sutcliffe, Carol A. Mathews, Evonne McArthur, Helen Hatfield, Dia Kabir, Evan J. Giangrande, Rebecca G. Fortgang, Shirley B. Wang, Rakesh Karmacharya, Joshua L. Roffman, Jeremiah M. Scharf, Jordan W. Smoller, Takahiro Soda, James J. Crowley, Lea K. Davis

## Abstract

**Objectives:** Obsessive-compulsive disorder (OCD) is a common psychiatric disorder, with two-thirds of affected individuals reporting severe impairment. Despite its substantial burden and moderate heritability, its etiology remains poorly understood, and treatments are often suboptimal. While recent genome-wide association studies (GWAS) have identified some risk loci, yet OCD remains in the linear phase of sample collection to variant association, with many more OCD-associated variants left to discover. This study aimed to develop and validate an electronic health record (EHR)-based algorithm to identify OCD cases and facilitate large-scale genetic studies.

**Methods:** We leveraged EHR-linked biobank data from two large hospital systems, namely Vanderbilt University Medical Center (VUMC) and Mass General Brigham (MGB), to develop a high-throughput phenotyping algorithm integrating diagnostic codes, medication records, and natural language processing (NLP) of clinical notes. Algorithm performance was evaluated through expert chart review, and genetic validation was performed using OCD polygenic risk scores (PRS).

**Results:** Expert chart reviews found that our algorithm combining both ICD codes and NLP achieved higher positive predictive values (PPV) for OCD cases (0.84 at VUMC and 0.91 at MGB) compared to using either ICD codes or NLP alone, albeit with a lower case yield.

Furthermore, at both sites, algorithm-determined cases exhibited significantly elevated PRS derived from the latest OCD GWAS, providing genetic validation of our phenotyping approach.

**Conclusion:** Our study demonstrates a scalable and cost-efficient approach for EHR-based ascertainment of OCD cases, facilitating large-scale genetic studies and advancing understanding of the disorder’s complex etiology.

## INTRODUCTION

Obsessive-compulsive disorder (OCD) is a common psychiatric disorder with a lifetime prevalence of 2.3% and an annual prevalence of 1.2% [1]. It is characterized by intrusive thoughts or impulses that typically cause anxiety or distress (obsessions), and repetitive mental acts or behaviors that one feels compelled to do (compulsions). OCD is highly debilitating; two-thirds of those affected report severe impairment, averaging 45 days of the past year spent outside normal roles at work, home, or socially [1]. Globally, OCD was once ranked the 10^th^ most disabling illness, as measured by years lived with disability (YLD), just behind schizophrenia, owing to its high prevalence, chronic course and pervasive symptoms [2]. Symptoms typically emerge early in life, with a mean onset at age 19 [1]. For a majority of individuals, OCD is a chronic condition, with remission achieved in only about one-fifth of cases [3]. Factors associated with lower remission rates include greater symptom severity [3,4], earlier age of onset [4], and longer illness duration [5].

Despite its significant burden, the etiology of OCD remains poorly understood, and existing treatments are suboptimal. Twin studies have consistently demonstrated higher concordance rates for OCD in monozygotic compared to dizygotic twins [6–10], with estimated heritability around 40-50% [9,11]. Early genome-wide association studies (GWAS) for OCD were limited by small sample sizes (<3,500 cases) [12,13] but established OCD as a complex genetic trait, indicating that larger sample sizes could yield significant loci. The most recent OCD GWAS identified 30 genome-wide significant loci [14]; however, the total genotyped sample size remains modest compared to other psychiatric disorders.

National disease registers, such as those in Nordic countries, have proven effective for large-scale sample collection. Diagnostic codes within these registers have generally shown high positive predictive values (PPV), around 90%. For instance, two register-based studies in Sweden and Denmark reported PPVs ranging from 85-96% [15,16]. Electronic health records (EHR) provide an alternative and efficient strategy for case ascertainment, successfully utilized in psychiatric research for bipolar disorder [17], treatment resistant depression [18], binge eating disorder [19], and autism spectrum disorder [20,21].

Existing OCD GWAS [12–14,22] have employed both family and unrelated sample designs. Notably, Strom et al. [14] conducted a large-scale GWAS meta-analysis of 28 OCD case-control cohorts of European ancestry, comprising cases from clinical settings, national registers, EHR-linked biobanks, and consumer self-report data. Their approach did not include rigorous phenotyping involving detailed clinical assessments or natural language processing (NLP) of clinical notes. Thus, comprehensive, multi-dimensional phenotyping integrating structured and unstructured EHR data remains underexplored for OCD.

In this study, we report the development and validation of a high-throughput phenotyping algorithm that integrates NLP, ICD codes, and medication administration records to systematically identify relevant evidence and determine patient OCD status across two academic medical centers: Vanderbilt University Medical Center (VUMC) and Mass General Brigham (MGB). Both institutions are part of PsycheMERGE, a network of EHR-linked biobanks for precision psychiatry research. Our algorithm was rigorously validated by expert chart review, demonstrating high PPV at both centers. Applying this method, we identified 417-1057 samples at VUMC and 335-1379 samples at MGB, varying by algorithmic definition, for DNA genotyping. Subsets of identified cases (of European genetic ancestry) exhibited significantly elevated polygenic risk scores (PRS) derived from the latest OCD GWAS [14]. Our results demonstrate that automated EHR-based phenotyping can effectively identify OCD cases and controls with precision, yielding samples genetically comparable to those ascertained through traditional, resource-intensive methods. This approach represents a valuable tool for accelerating psychiatric genetic research.

## METHODS

### Study participants

This research was conducted as part of the Psychiatric Genomics Consortium’s (PGC) ongoing effort to identify and collect DNA samples from patients with OCD, using two EHR-linked biobanks within the PsycheMERGE Network: BioVU at VUMC in Nashville, Tennessee, and the MGB Biobank (MGBB) [23] at MGB in Boston, Massachusetts. The study included 213,275 individuals receiving care at VUMC from 1989 to 2021, and 97,275 patients at MGB from 1976 to 2021. Ethical approvals were obtained from the institutional review boards at both VUMC and MGB, including an informed consent waiver for the use of retrospective medical record data without patient interaction. All participants provided written informed consent for their inclusion in the respective biobanks for broad-based research use. Additional data details are provided in Appendix S1 (Supplementary Material).

### OCD phenotyping algorithm

Our OCD algorithm was developed at VUMC through an iterative, consensus-driven process, involving domain experts from PGC, VUMC psychiatry, and neurology. Relevant ICD-9 and ICD-10 codes for inclusion and exclusion (Table S3 and S4) were selected for the structured data components of the rule-based algorithm, while diagnostic and treatment-related keywords were incorporated into its NLP arm (Table S5). To establish a broad performance base, we included ICD codes for body dysmorphic disorder (BDD) and trichotillomania (TTM), with the understanding that these criteria can be modified to adjust the algorithm’s specificity for different use cases. Medications were extracted from multiple sources, including structured EHR databases containing recorded prescriptions and unstructured clinical notes. Once all algorithm components were finalized and the logical order of its operations was confirmed by the domain experts, we implemented the algorithm within the VUMC clinical database. Three rounds of chart review were conducted at VUMC and MGB, respectively, to validate its performance. Detailed steps in developing the algorithm are provided in Appendix S2.

The resulting algorithm integrates diagnostic, medication, and clinical note data to determine each patient’s OCD status. It begins with exclusion criteria (Figure 1A), followed by two inclusion pathways: (1) an ICD arm, which identifies patients based on ICD-9 and ICD-10 codes for OCD, BDD and TTM (Figure 1B); and (2) an NLP arm, detecting OCD-related mentions in patients’ clinical notes (Figure 1C). Exclusion criteria include ICD codes for “Other specified metabolic disorders” and “Metabolic disorder, unspecified” to prevent rare but highly penetrant metabolic disorders, such as Lesch-Nyhan syndrome, from being misidentified as OCD. We derived four definitions of OCD from this algorithm: (1) ICD-defined, including cases identified exclusively through the ICD arm of the algorithm; (2) NLP-only, encompassing cases that lacked ICD-coded information but are identified through the NLP arm based on clinical notes; (3) ICD-or-NLP, identifying cases from either the ICD or the NLP arm, i.e., the disjunction of the two arms resulting in the union of cases; and (4) ICD-and-NLP, requiring concordance between the two arms, i.e., the conjunction of both arms representing the intersection of cases. We used the ICD-or-NLP definition to sample OCD cases for chart reviews, as it represents the broadest and most inclusive version of our algorithm.

**Figure 1:**
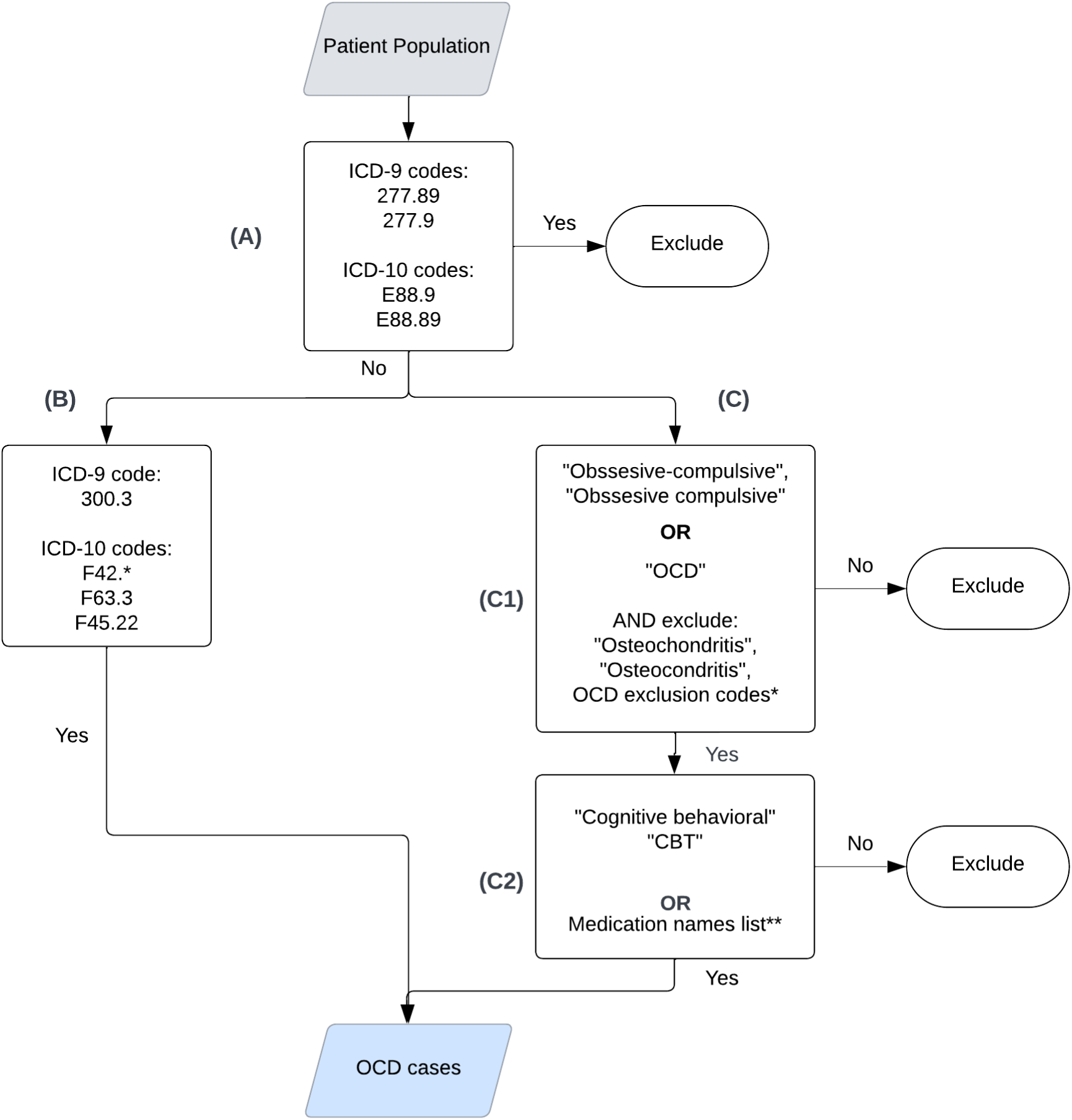
OCD phenotyping algorithm flowchart at VUMC and MGB, consisting of an ICD codes-based exclusion criteria (A), an ICD codes-based inclusion arm (B), and a rule-based NLP algorithm arm (C). *ICD codes for OCD exclusion: Table S4 (Supplementary Material). **Medication names list: Appendix Table S5 (Supplementary Material).

#### NLP system

We developed and integrated a rule-based NLP system into the algorithm to identify OCD mentions from clinical notes (Figure 1C), comprising two steps: (1) identifying OCD keyword mentions (C1); and (2) identifying cognitive behavioral therapy (CBT) keyword mentions and relevant medications (C2). To meet the criteria in C1, a patient must have at least two mentions of OCD on separate days in their hospital notes, such as problem lists, discharge summaries, clinical communications, progress notes, or visit notes, with the first OCD mention before age 55 [24,25]. This requirement accounts for the possibility that late-life OCD diagnosis may more likely reflect changes due to cognitive decline, often with a biological etiology distinct from that of idiopathic OCD. We also incorporated exclusion criteria in C1, using keywords and codes, to disambiguate the acronym “OCD” and prevent false identifications, such as mentions related to osteochondritis dissecans. In C2, any mention of OCD treatments including CBT or relevant medications are identified and considered as additional evidence for OCD. Medications were extracted from two sources: (a), the structured patient medication database; and (b) unstructured clinical notes, using the MedEx-UIMA system [26], based on an OCD medication list (Table S5). Further details about the NLP system are provided in Appendix S3.

### Chart review

To evaluate the accuracy of our algorithm, two clinically trained reviewers (EM: a medical student post-psychiatry clerkship; PM: a psychiatry resident) conducted manual chart reviews at VUMC. All the sampled OCD cases for chart review were identified using ICD-or-NLP. Both reviewers independently examined charts from the same set of 25 subjects (including 5 subjects not identified by the algorithm as OCD cases), to calculate inter-rater reliability (IRR). Reviewers were blinded to each other’s evaluation. For each subject, reviewers determined the true OCD diagnosis and flagged the level of supporting evidence. Two psychiatrists (JB, HH) then reviewed these findings, provided feedback, and adjudicated discrepancies. Once adequate agreement (IRR>90%) was achieved, each reviewer independently assessed additional 25 subjects. If agreement was inadequate, another 25 subjects were reviewed, followed by the same confirmation process until the required agreement threshold was met. Once confirmed, the remaining subjects were reviewed independently. Detailed reviewer instructions are provided in Appendix S4.

For external validation at MGB, two board-certified psychiatrists (JR, RK) conducted two rounds of chart reviews for algorithm tuning and evaluation. Any disagreements between the two reviewers were adjudicated by a third psychiatrist (JS). In the first round, 23 algorithm-identified cases and three algorithm-identified non-cases (by ICD-or-NLP) were randomly sampled. A second round involved 45 cases and five non-cases randomly sampled. Finally, 90 algorithm-identified cases (by ICD-or-NLP) were randomly sampled and reviewed by three clinical psychologists (EG, RF, SW) who initially reviewed the same set of 10 subjects for training and calibration. Each chart was subsequently reviewed by two reviewers, with disagreements adjudicated by the third reviewer, resulting in each reviewer evaluating 33-35 charts.

### Genetic Validation (VUMC)

In 2007, VUMC launched BioVU, a biobank linking patient DNA samples with EHRs. The BioVU consent form is provided to patients at outpatient clinic settings. We applied quality control measures (described in Appendix S5.1) to the algorithm-determined OCD cases and controls in BioVU to generate the final dataset used for genetic validation.

Polygenic risk scores (PRS) were calculated based on the GWAS conducted by Strom et al. [14], which included 22,479 OCD cases and 1,076,000 controls after excluding 23andMe and BioVU samples. We employed PRS-CS [27] to estimate posterior SNP effect sizes, using CEU (refers to Utah residents with Northern and Western European ancestry) samples from the 1000 Genomes Project Phase 3 as the reference panel. PRS was computed as the sum of the risk allele dosages weighted by the posterior SNP effect sizes. Logistic regression analyses were conducted to assess the association between OCD PRS and the algorithm-derived OCD status, adjusting for current age, median age of medical record, sex, genotype batch, and principal component analysis (PCA) components.

### Genetic Validation (MGB)

The MGB Biobank (MGBB), established in 2010, collects biological samples, health information, and genetic data to support biomedical research [23]. Participants are enrolled using a broad-based consent process by research coordinators at clinical and public hospital locations or electronically through the MGB patient portal. Genotyping was performed for 53,853 participants in 13 batches using four Illumina array platforms: Multi-Ethnic Genotyping Array (MEGA, batch 1), Expanded Multi-Ethnic Genotyping Array (MEGAEX, batch 2), Multi-Ethnic Global (MEG, batches 3–9), and Global Screening Array (GSA, batches 10–13). Samples from batches 8 and 9 were excluded due to a limited number of OCD cases identified (15 and 10 cases, respectively, using ICD or NLP criteria). Quality control procedures applied in genetic validation are detailed in Appendix S5.2.

Consistent with methods employed at VUMC, SNP effect sizes for OCD were obtained from Strom et al’s GWAS [14] without 23andMe individuals. Posterior SNP effect sizes were calculated using PRS-CS [27], referencing CEU samples from the 1000 Genomes Project Phase 3. PRS values were calculated as the weighted sum of risk allele dosages, using the computed posterior SNP effect sizes. Logistic regression analyses were conducted separately in the MEG and GSA datasets to evaluate associations between OCD PRS and the algorithm-determined OCD status, adjusting for age and PCA components.

## RESULTS

### Chart review validation

Three rounds of chart review were conducted at VUMC. In the first two rounds, 50 charts were reviewed by two reviewers, achieving a raw agreement of 0.91 and a Cohen’s kappa of 0.72, indicating substantial IRR [28]. In the third and final round, each reviewer independently reviewed 22-25 charts, bringing the total number of charts reviewed to 97. At MGB, two initial rounds of chart review identified 34 OCD cases and 42 non-cases, with the reviewers reaching the same conclusion on 71 out of 76 charts (raw agreement=0.93; Cohen’s kappa=0.91). For the remaining five charts, a third reviewer provided the final decision. In the final round at MGB, three reviewers assessed 90 charts (52 OCD cases and 38 non-cases), achieving raw agreement of 0.87 and Cohen’s kappa of 0.71. At MGB, only the final round was used for algorithm validation (Table 1).

**Table 1.** Comparison of the four OCD definitions in terms of PPV and the number of genotyped OCD cases identified in the biobanks. ICD-defined refers to charts selected based on ICD-coded information regardless of whether NLP was positive. NLP-only refers to charts that lacked ICD-coded information but were algorithm positive based on NLP applied to unstructured notes. ICD-or-NLP: uses both arms and takes the union of the cases identified by either arm. ICD-and-NLP: uses both arms and takes the intersection of cases identified by both arms.

| Algorithmic definition | VUMC |  | MGB |  |
| --- | --- | --- | --- | --- |
|  | PPV | N genotyped | PPV | N genotyped |
| ICD-or-NLP | 0.72 | 1057 | 0.58 | 1379 |
| ICD-defined | 0.73 | 640 | 0.64 | 725 |
| NLP-only | 0.71 | 417 | 0.51 | 654 |
| ICD-and-NLP | 0.84 | 515 | 0.91 | 335 |

Table 1 presents the chart review validation results from both sites. Using the intersection of cases from both arms (ICD-and-NLP) yielded the highest positive predictive values (PPV) of 0.84 at VUMC and 0.91 at MGB, compared to using the union (ICD-or-NLP) or individual algorithm arms (ICD-defined, NLP-only). However, ICD-and-NLP identified fewer ICD cases (515 at VUMC and 335 at MGB). Conversely, ICD-or-NLP identified the highest number of cases (1057 at VUMC and 1379 at MGB) but resulted in lower PPVs of 0.72 at VUMC and 0.58 at MGB, with 38 out of 90 subjects at MGB misclassified as OCD cases.

These findings highlight the tradeoff between PPV and the number of identified OCD cases when comparing the disjunction (ICD-or-NLP) and conjunction (ICD-and-NLP) of the two arms in the algorithm. The NLP-only definition, which includes only cases identified by the NLP arm and excludes cases with relevant ICD codes, yielded the lowest PPVs at both sites. Replication at MGB resulted in significantly lower PPV in three of the four definitions (ICD-defined, NLP-only, and ICD-or-NLP), most notably in NLP-only and ICD-or-NLP, underscoring the challenges associated with using clinical notes across different sites.

### Description of algorithm results

Among the 213,275 BioVU participants, our algorithm identified 640 genotyped OCD cases using the ICD arm alone (ICD-defined), 417 cases exclusively via the NLP arm (NLP-only) who had no relevant ICD codes, 1,057 cases through either arm (ICD-or-NLP), and 515 cases identified concordantly by both arms (ICD-and-NLP). For MGBB, 3,385 of 97,275 participants were excluded based on predefined criteria (Figure 1A). Of the remaining 93,890 participants, 53,853 underwent genotyping, yielding 725 cases by ICD-defined, 654 by NLP-only, 1,379 by ICD-or-NLP, and 335 by ICD-and-NLP. Pie charts summarizing the OCD case counts are presented in Figure S3.

Table 2 and S2 (Supplementary Material) summarize the demographic characteristics of the algorithm-identified OCD cohorts at VUMC and MGB. Cohorts identified solely through the ICD arm had a higher proportion of patients with public health insurance (55.16% at VUMC; 65.38% at MGB) and, on average, older patients (46.22 years at VUMC; 55.35 years at MGB) compared to other cohorts. In contrast, NLP-only cohorts had a lower proportion of patients with public insurance (45.56% at VUMC; 50.76% at MGB) and younger average age (43.99 years at VUMC; 45.17 years at MGB). All cohorts had greater female representation than male, particularly in the NLP-only cohorts (65.71% at VUMC; 67.28% at MGB). Most individuals across cohorts self-reported as White (91.10%-92.43% at VUMC; 82.72%-90.75% at MGB), with the ICD-and-NLP cohorts having the highest proportions. In terms of hospital utilization, the NLP-only cohorts consistently had the lowest counts for visits, diagnoses (ICD), procedures (CPT), prescriptions (RxNorm), lab tests (LOINC), and clinical notes across both sites. The cohort with the highest utilization varied, being the ICD-and-NLP cohort at VUMC and the ICD-only cohort at MGB. Notably, despite lower overall utilization and a shorter study time frame (1989-2021 at VUMC vs. 1976-2021 at MGB), cohorts at VUMC had markedly higher clinical note counts, suggesting differences in documentation practices between the two sites.

**Table 2.** Demographic composition of the OCD cases identified by each arm of the algorithm.

| Setting | ICD-defined<br>(VUMC) | NLP-only<br>(VUMC) | ICD-defined<br>(MGB) | NLP-only<br>(MGB) |
| --- | --- | --- | --- | --- |
| Genotyped *N* | 640 | 417 | 725 | 654 |
| Age, mean (SD) | 46.22 (17.53) | 43.99 (15.61) | 55.35 (17.58) | 45.17 (11.03) |
| Gender: *N* (%) |  |  |  |  |
| *Female* | 357 (55.78) | 274 (65.71) | 442 (60.97) | 440 (67.28) |
| *Male* | 283 (44.22) | 143 (34.29) | 283 (39.03) | 214 (32.72) |
| Self-reported<br>Race: *N* (%) |  |  |  |  |
| *Asian* | 9 (1.41) | 4 (0.96) | 10 (1.38) | 17 (2.60) |
| *Black/African*<br>*American* | 36 (5.63) | 21 (5.04) | 29 (4.0) | 33 (5.05) |
| *White* | 583 (91.10) | 382 (91.61) | 646 (89.10) | 541 (82.72) |
| *Other* | 3 (0.47) | 1 (0.24) | 30 (4.14) | 54 (8.26) |
| *Unknown* | 9 (1.41) | 9 (2.16) | 10 (1.38) | 9 (1.38) |
| Ethnicity: *N* (%) |  |  |  |  |
| *Hispanic* | 11 (1.72) | 12 (2.88) | 12 (1.66) | 20 (3.06) |
| *Non-Hispanic* | 629 (98.28) | 405 (97.12) | 713 (98.34) | 634 (96.94) |
| Public payer: *N*<br>(%) |  |  |  |  |
| *Yes* | 353 (55.16) | 190 (45.56) | 474 (65.38) | 332 (50.76) |
| *No* | 287 (44.84) | 227 (54.44) | 251 (34.62) | 322 (49.24) |
| Hospital usage: Median (IQR) |  |  |  |  |
| *EHR length (in* | 7.78 (11.98) | 8.53 (12.26) | 18.47 (11.62) | 14.50 (13.03) |
| *years)* |  |  |  |  |
| *Visit count* | 76 (127.25) | 63 (105) | 366 (463.0) | 268.5 (362.75) |
| *ICD count* | 208.5 (408.75) | 167 (324) | 809 (1271.0) | 587.5 (809.0) |
| *CPT count* | 215 (339.25) | 167 (298) | 593 (867.0) | 405 (595.25) |
| *RxNorm count* | 117.5 (158.25) | 99 (138.75) | 566.0 (948.0) | 390.0 (696.75) |
| *LOINC count* | 159 (129.5) | 139.5 (127) | 525.5 (532.75) | 442.0 (457.75) |
| *Note count* | 1155 (2255) | 1081 (2146.75) | 363 (544.0) | 273.5 (405.25) |

### Genetic validation

#### VUMC

The OCD PRS was calculated for individuals of European genetic ancestry in the VUMC BioVU cohort using the OCD GWAS reported by Strom et al. [14], which included 53,660 OCD cases and 2,044,417 controls of European ancestry. A total of 5,083,270 SNPs overlapped between the OCD GWAS and the BioVU dataset, with posterior SNP effect sizes were estimated for 769,851 SNPs using PRS-CS. OCD PRS of BioVU individuals was calculated as the sum of the risk allele weighed by their posterior SNP effect sizes. Genetic validation analyses were restricted to individuals of European ancestry labeled as OCD cases and controls by each algorithmic definition, for example: ICD-or-NLP (cases = 676, controls = 46,677) and ICD-and-NLP (cases = 326, controls = 46,677).

Using logistic regression, a significant association between OCD PRS and predicted OCD status using the ICD-or-NLP definition was observed in BioVU (OR = 1.21, SE = 0.039, P = 9.48E-07). A significant association was also found using the ICD-and-NLP definition, with a higher effect size but less significant p-value (OR = 1.27, SE = 0.056, P = 1.61E-05), likely due to the smaller case size.

**Table 3.** Association between OCD PRS and predicted OCD status in VUMC BioVU. OR: Odds ratio. BETA: Beta coefficient. SE: Standard error. P: P-value. R^2^: Pseudo-R-squared by Nagelkerke.

| Algorithmic definition | Dataset | N of cases | N of controls | OR | BETA | SE | P | R<sup>2</sup> |
| --- | --- | --- | --- | --- | --- | --- | --- | --- |
| ICD-defined | BioVU | 398 | 46,677 | 1.25 | 0.221 | 0.051 | 1.47E-05 | 0.86% |
| NLP-only | BioVU | 278 | 46,677 | 1.16 | 0.151 | 0.061 | 0.013 | 0.68% |
| ICD-or-NLP | BioVU | 676 | 46,677 | 1.21 | 0.193 | 0.039 | 9.48E-07 | 1.50% |
| ICD-and-NLP | BioVU | 326 | 46,677 | 1.27 | 0.242 | 0.056 | 1.61E-05 | 0.76% |

#### MGB

A total of 1,379 OCD cases (ICD-or-NLP) and 52,474 controls were genotyped in 13 batches on 4 Illumina genotyping platforms. After quality control, duplicated or related samples, non-European samples, and samples and SNPs with low genotyping quality were removed. This left 658 cases and 23,314 controls with 593,590 SNPs in the MEG dataset, and 388 cases and 13,458 controls with 407,608 SNPs in the GSA dataset. After matching controls to cases at a 4:1 ratio based on gender, genotyping batch, age, and genetic proximity, imputation was performed separately for MEG (658 cases, 2,632 controls) and GSA (388 cases, 1,552 controls). A total of 5,866,159 SNPs passed post-imputation quality control in both datasets.

PRS calculations for MGB were also based on the OCD GWAS [14]. A total of 5,598,712 SNPs overlapped between the OCD GWAS and the MGBB datasets, and posterior SNP effect sizes were estimated for 1,048,347 SNPs using PRS-CS. PRS of MGBB individuals was also calculated as the sum of the risk allele dosages weighed by their posterior SNP effect sizes. Logistic regression analyses, adjusting for population stratification, found significant associations between standardized PRS and predicted OCD status using the ICD-or-NLP definition in both MEG and GSA (OR = 1.13, SE = 0.044, P = 0.0051; OR = 1.31, SE = 0.058, P = 4.3×10^-6^, respectively). Similar associations were observed using the ICD-and-NLP definition, with higher effect sizes but less significant p-values (MEG: OR = 1.25, SE = 0.089, P = 0.011; GSA: OR = 1.39, SE = 0.126, P = 0.0095), likely due to the smaller sample size. Meta-analysis across two datasets yielded ORs of 1.19 (P = 5.4×10^-7^) for ICD-or-NLP and 1.30 (P = 3.7×10^-4^) for ICD-and-NLP. No significant association was observed using the NLP-only definition (OR = 1.08, SE = 0.052, P = 0.13), consistent with its lower PPV at MGB as reported in Table 1.

**Table 4.**
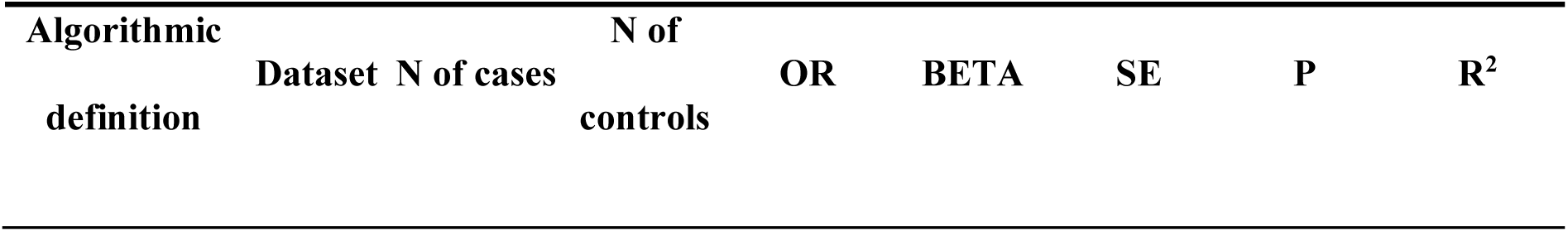
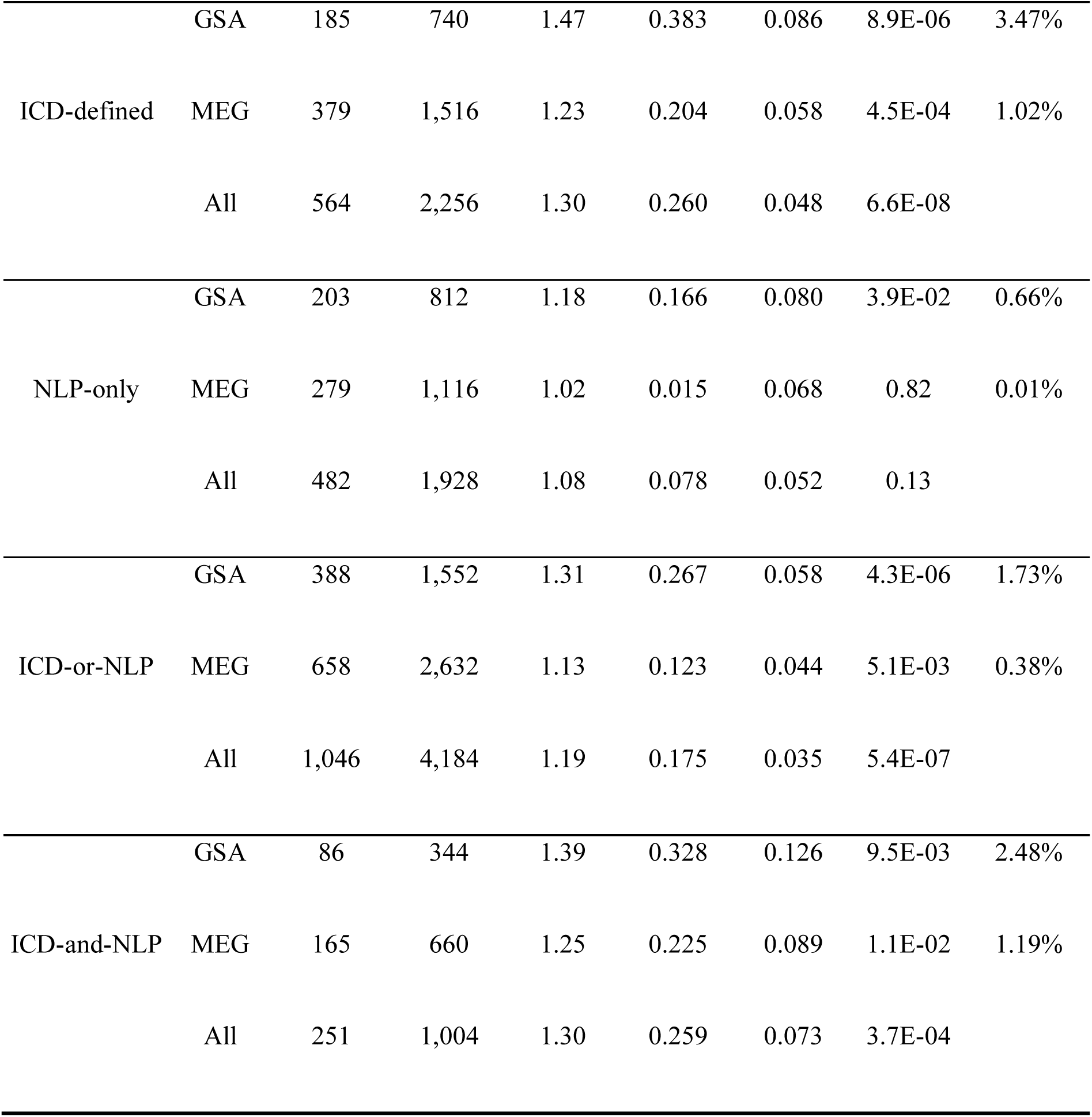
Association between OCD PRS and predicted OCD status in MGBB. OR: Odds ratio. BETA: Beta coefficient. SE: Standard error. P: P-value. R^2^: Pseudo-R-squared by Nagelkerke.

## DISCUSSION

OCD is a common psychiatric condition, yet remission is achieved in only about a fifth of cases [3]. Unfortunately, the interval between symptom onset and treatment initiation averages 7 years [29], and longer duration of untreated OCD is associated with poorer outcomes [30]. Given this, identifying biological markers and genetic risk factors is crucial for facilitating earlier diagnosis and targeted interventions. Large scale agnostic gene discovery studies of OCD are gaining traction and recent work demonstrates discovery yield increases as sample size scales upwards [14]. Previous OCD GWAS have relied on cases ascertained from various sources, including specialist clinics and advocacy organizations [12,13], or national disease registers from the Nordic countries [15,16]. The availability of large-scale, real-world EHR data has provided an opportunity to develop scalable and cost-efficient tools to identify patients with psychiatric disorders, such as OCD, for genetic studies. A recent study utilized EHR data from a mental health case register in the UK to ascertain obsessive-compulsive symptoms (OCS) and OCD from structured fields and free-text records processed via NLP; however, no external evaluation was conducted at a geographically distinct site, leaving the question of algorithm generalizability open [31].

In this study, we developed a high-throughput EHR-based algorithm for identifying OCD, comprising an ICD arm that utilizes ICD-9 and ICD-10 codes, and an NLP arm that incorporates medication records and applies NLP to clinical notes. Our algorithm demonstrated high PPV compared to multiple rounds of clinical chart reviews, showing generalizability across two healthcare systems, VUMC and MGB, located in the southeastern and northeastern United States. Leveraging recent OCD GWAS data from Strom et al. [14], we calculated PRS for individuals of European genetic ancestry in our VUMC BioVU and MGBB cohorts and found significant associations between OCD PRS and the algorithm-determined OCD status via logistic regression. This provides early evidence that our EHR-based algorithm can be used to ascertain OCD cases that are genetically comparable to traditionally ascertained samples, providing substantial time and cost efficiencies in assembling cohorts for genetic research. As outlined below, three key findings from our analyses are particularly noteworthy.

First, our expert chart reviews (Table 1) showed that using the intersection of cases of both algorithm arms (ICD-and-NLP) achieved significantly higher PPVs (0.84 at VUMC and 0.91 at MGB) than using either arm individually (ICD-defined, NLP-only) or their union (ICD-or-NLP) at both sites, particularly at MGB, where the PPVs for each individual arm were only 0.64 and 0.51. These findings suggest the ICD codes and clinical notes processed via NLP are complementary in enhancing the precision of ascertaining OCD phenotypes from EHR data when used together. On the other hand, ICD-and-NLP also resulted in the lowest number of identified OCD cases (515 at VUMC, 335 at MGB) for inclusion in the downstream genetic studies, whereas ICD-or-NLP identified the most cases (1057 at VUMC, 1379 at MGB) but with substantially lower PPVs (0.72 at VUMC, 0.58 at MGB). This underscores that the two arms are complementary only when used in conjunction since the union of their identified cases (disjunction) tends to accumulate false positives, resulting in poorer PPVs than when each arm is applied independently. Moreover, this pattern of findings highlights the tradeoff between achieving high precision (PPV) and maximizing the number of cases identified when selecting an algorithmic definition. Additionally, we observed PPVs achieved by using only ICD codes to select cases at the two U.S.-based healthcare systems (PPV = 0.73 and 0.64) were notably lower than those reported in studies from the Nordic countries (0.91–0.96 in Sweden [15], 0.85–0.96 in Denmark [16]), suggesting less reliability of ICD-only ascertainment in the U.S. compared to Nordic registers.

Second, among individuals of European ancestry, we observed significant associations between OCD PRS derived from Strom et al.’s GWAS and OCD phenotypes identified by our algorithm definitions, including ICD-or-NLP (OR = 1.21, SE = 0.039, P = 9.48E-07 at VUMC; OR = 1.19, SE = 0.035, P = 5.4E-07 at MGBB) and ICD-and-NLP (OR = 1.27, SE = 0.056, P = 1.61E-05 at VUMC; OR = 1.33, SE = 0.074, P = 1.3E-04 at MGBB). This provides early evidence that the proposed algorithm captures genetic influences that overlap with those observed in OCD samples identified through traditional ascertainment methods. In addition to further validating the genetic basis of our EHR-derived phenotypes, these results suggest that such samples can be integrated with traditional cohorts to enhance the statistical power of genetic discovery. Furthermore, we observed that when used in isolation, the NLP arm produced the smallest effect sizes (with the least significant p-values), emphasizing challenges in relying solely on rule-based NLP for ascertaining OCD cases from clinical narratives.

Finally, replication and validation of our algorithm across two healthcare systems provided several key insights: (1) variability in local documentation practices and clinical workflows can affect data completeness and complicates algorithm replication; (2) standardized chart review processes, including consistent use of reviewer training materials (Appendix S4), helped ensure reproducibility of chart review results across sites; (3) open communication between teams across sites facilitated harmonization of algorithm development and troubleshooting during replication; (4) comparing chart reviews between VUMC and MGB revealed site-specific variations in PPV, notably improved PPV for ICD-and-NLP at MGB (from 0.84 to 0.91), but reductions in PPV for other definitions, especially NLP-only (from 0.71 to 0.51). On one hand, this shows the heterogeneity of clinical notes, and the challenge associated with processing unstructured clinical text across healthcare systems. On the other hand, these results also suggest that using both the ICD and NLP arms in conjunction may help mitigate such challenges, including variability in coding practices and limited transferability of rule-based NLP models; (5) the convergent validation of OCD cases via genetic evidence, demonstrated through significant associations with OCD PRS derived from GWAS, provided an important external benchmark for measuring phenotype accuracy beyond standard chart reviews. Together, these findings highlight the importance of developing flexible yet standardized phenotyping methods, accounting for site-specific data differences, and employing consistent validation strategies to strengthen confidence in algorithm reproducibility and generalizability.

There are several limitations of the presented study: (1) cross-site validation of our OCD phenotyping algorithm is complicated by variations in data availability and completeness. Discrepancies often arise between when diagnostic codes are recorded and when corresponding clinical notes are accessible, in part because notes may be sequestered or otherwise unavailable, whereas codes remain accessible. This issue is particularly pronounced for psychiatric notes, which have historically had restricted access due to privacy safeguards. Although recent changes under the ‘Open Notes’ mandate of the 21st Century Cures Act have begun to reduce these barriers, adoption remains inconsistent across institutions, resulting in persistent data gaps due to administrative, technical, or privacy constraints. Moreover, since patients often receive care across multiple systems, only partial clinical documentation or diagnostic codes may appear within any single site’s EHR, further complicating algorithm replication; (2) rule-based NLP models, including ours, often fail to fully grasp the context of medical concepts in clinical notes and struggle with semantic ambiguity due to their reliance on rigid, hand-crafted rules. Our analysis at MGB demonstrated this limitation through specific examples: the NLP arm of the algorithm incorrectly identified cases by missing negations (such as ‘no’ responses in poorly formatted questionnaires), failing to recognize copy-pasted content from templates, and misinterpreting abbreviations such as “OCD”, confusing obsessive-compulsive disorder with osteochondritis dissecans; (3) the OCD PRS was calculated only for individuals of European genetic ancestry in our study cohorts due to availability of existing GWAS summary statistics. As a result, our findings may not generalize well to populations with different genetic ancestral backgrounds.

Our current work within the PsycheMERGE network is aimed at establishing a standardized review protocol by training chart reviewers across multiple healthcare institutions using simulated psychiatric patient records. This standardization and calibration aim to directly address the reduced generalizability of phenotyping algorithms due to inconsistencies in chart review strategies. Recent advances in large language models (LLMs) offer an opportunity to develop phenotyping algorithms in a highly scalable way without substantial annotated data. Future work will explore using LLMs to identify OCD-related information in clinical notes, supported by recent findings demonstrating superior diagnostic accuracy of LLMs compared to mental health professionals in identifying OCD cases from clinical vignettes [32]. Furthermore, we will diversify the genetic ancestry representation in our OCD validation analyses by utilizing resources such as the All of Us Research Program [33]. This will allow us to develop more inclusive and equitable models of OCD genetic risk that better represent global populations, directly addressing the limitations of current European-centric genetic studies.

## CONCLUSION

In conclusion, we developed and validated an EHR-based phenotyping algorithm for OCD integrating diagnostic codes, medication records, and unstructured clinical notes across two healthcare systems (VUMC and MGB). Our findings demonstrate that combining the ICD and NLP arms of this algorithm can identify OCD cases with high precision, yielding cohorts that present significant associations with OCD PRS derived from GWAS at both sites. This high-throughput, cost-efficient approach can facilitate large-scale genetic studies and advance understanding of OCD’s complex etiology.

## ACKNOWLEDGEMENTS

We would like to acknowledge Drs. Patrick McGuire and Jonathan Becker for their contribution to this study as clinical chart reviewers.

## FUNDING

LKD and DH were supported by R01MH137220 and R01MH118233 awarded from the National Institute of Mental Health. The Synthetic Derivative and BioVU projects at VUMC are supported by numerous sources: including the NIH funded Shared Instrumentation Grant S10OD017985 and S10RR025141; CTSA grants UL1TR002243, UL1TR000445, and UL1RR024975 from the National Center for Advancing Translational Sciences. Its contents are solely the responsibility of the authors and do not necessarily represent official views of the National Center for Advancing Translational Sciences or the National Institutes of Health. Genomic data are also supported by investigator-led projects that include U01HG004798, R01NS032830, RC2GM092618, P50GM115305, U01HG006378, U19HL065962, R01HD074711; and additional funding sources listed at https://victr.vumc.org/biovu-funding/.

## CONFLICT OF INTEREST

Dr Smoller reported grants from Biogen, Inc and serving as a scientific advisory board member with options from Sensorium Therapeutics, Inc outside the submitted work. Dr Wang reported receiving grants from the Brain and Behavior Research Foundation during the conduct of the study. No other disclosures were reported.

## DATA AVAILABILITY

Protected Health Information restrictions apply to the availability of the clinical data here, which were used under IRB approval for use only in the current study. As a result, this dataset is not publicly available.

## Notes

### Author Declarations

Ethical approval was obtained from the institutional review boards at both Vanderbilt University Medical Center and Mass General Brigham.

## REFERENCES

1 Ruscio AM, Stein DJ, Chiu WT, et al. The epidemiology of obsessive-compulsive disorder in the National Comorbidity Survey Replication. Mol Psychiatry 2010;15:53–63. doi:10.1038/mp.2008.94

2 Lopez AD, Murray CC. The global burden of disease, 1990-2020. Nat Med 1998;4:1241–3. doi:10.1038/3218

3 Law C, Kamarsu S, Obisie-Orlu IC, et al. Personality traits as predictors of OCD remission: A longitudinal study. J Affect Disord 2023;320:196–200. doi:10.1016/j.jad.2022.09.121

4 Geiger Y, van Oppen P, Visser H, et al. Long-term remission rates and trajectory predictors in obsessive-compulsive disorder: Findings from a six-year naturalistic longitudinal cohort study. J Affect Disord 2024;350:877–86. doi:10.1016/j.jad.2024.01.155

5 Eisen JL, Sibrava NJ, Boisseau CL, et al. Five-year course of obsessive-compulsive disorder: predictors of remission and relapse. J Clin Psychiatry 2013;74:233–9. doi:10.4088/JCP.12m07657

6 Inouye E. Similar and dissimilar manifestations of obsessive-compulsive neurosis in monozygotic twins. Am J Psychiatry 1965;121:1171–5. doi:10.1176/ajp.121.12.1171

7 Clifford CA, Murray RM, Fulker DW. Genetic and environmental influences on obsessional traits and symptoms. Psychol Med 1984;14:791–800. doi:10.1017/s0033291700019760

8 Jonnal AH, Gardner CO, Prescott CA, et al. Obsessive and compulsive symptoms in a general population sample of female twins. Am J Med Genet 2000;96:791–6.

9 Mataix-Cols D, Boman M, Monzani B, et al. Population-based, multigenerational family clustering study of obsessive-compulsive disorder. JAMA Psychiatry 2013;70:709–17. doi:10.1001/jamapsychiatry.2013.3

10 Mataix-Cols D, Fernández de la Cruz L, Beucke JC, et al. Heritability of Clinically Diagnosed Obsessive-Compulsive Disorder Among Twins. JAMA Psychiatry 2024;81:631–2. doi:10.1001/jamapsychiatry.2024.0299

11 Davis LK, Yu D, Keenan CL, et al. Partitioning the heritability of Tourette syndrome and obsessive compulsive disorder reveals differences in genetic architecture. PLoS Genet 2013;9:e1003864. doi:10.1371/journal.pgen.1003864

12 Stewart SE, Yu D, Scharf JM, et al. Genome-wide association study of obsessive-compulsive disorder. Mol Psychiatry 2013;18:788–98. doi:10.1038/mp.2012.85

13 Mattheisen M, Samuels JF, Wang Y, et al. Genome-wide association study in obsessive-compulsive disorder: results from the OCGAS. Mol Psychiatry 2015;20:337–44. doi:10.1038/mp.2014.43

14 Strom NI, Gerring ZF, Galimberti M, et al. Genome-wide analyses identify 30 loci associated with obsessive–compulsive disorder. Nat Genet Published Online First: 13 May 2025. doi:10.1038/s41588-025-02189-z

15 Rück C, Larsson KJ, Lind K, et al. Validity and reliability of chronic tic disorder and obsessive-compulsive disorder diagnoses in the Swedish National Patient Register. BMJ Open 2015;5:e007520. doi:10.1136/bmjopen-2014-007520

16 Nissen J, Powell S, Koch SV, et al. Diagnostic validity of early-onset obsessive-compulsive disorder in the Danish Psychiatric Central Register: findings from a cohort sample. BMJ Open 2017;7:e017172. doi:10.1136/bmjopen-2017-017172

17 Castro VM, Minnier J, Murphy SN, et al. Validation of electronic health record phenotyping of bipolar disorder cases and controls. Am J Psychiatry 2015;172:363–72. doi:10.1176/appi.ajp.2014.14030423

18 Perlis RH, Iosifescu DV, Castro VM, et al. Using electronic medical records to enable large-scale studies in psychiatry: treatment resistant depression as a model. Psychol Med 2012;42:41–50. doi:10.1017/S0033291711000997

19 Bellows BK, LaFleur J, Kamauu AWC, et al. Automated identification of patients with a diagnosis of binge eating disorder from narrative electronic health records. J Am Med Inform Assoc 2014;21:e163–8. doi:10.1136/amiajnl-2013-001859

20 Bush RA, Connelly CD, Pérez A, et al. Extracting autism spectrum disorder data from the electronic health record. Appl Clin Inform 2017;8:731–41. doi:10.4338/ACI-2017-02-RA-0029

21 Malow BA, Veatch OJ, Niu X, et al. A practical approach to identifying autistic adults within the electronic health record. Autism Res 2023;16:52–65. doi:10.1002/aur.2849

22 International Obsessive Compulsive Disorder Foundation Genetics Collaborative (IOCDF-GC) and OCD Collaborative Genetics Association Studies (OCGAS). Revealing the complex genetic architecture of obsessive-compulsive disorder using meta-analysis. Mol Psychiatry 2018;23:1181–8. doi:10.1038/mp.2017.154

23 Karlson EW, Boutin NT, Hoffnagle AG, et al. Building the partners healthcare biobank at partners personalized medicine: informed consent, return of research results, recruitment lessons and operational considerations. J Pers Med 2016;6. doi:10.3390/jpm6010002

24 Frydman I, Ferreira-Garcia R, Borges MC, et al. Dementia developing in late-onset and treatment-refractory obsessive-compulsive disorder. Cogn Behav Neurol 2010;23:205–8. doi:10.1097/WNN.0b013e3181e61ce0

25 Frileux S, Millet B, Fossati P. Late-Onset OCD as a Potential Harbinger of Dementia With Lewy Bodies: A Report of Two Cases. Front Psychiatry 2020;11:554. doi:10.3389/fpsyt.2020.00554

26 Jiang M, Wu Y, Shah A, et al. Extracting and standardizing medication information in clinical text - the MedEx-UIMA system. AMIA Jt Summits Transl Sci Proc 2014;2014:37–42.

27 Ge T, Chen C-Y, Ni Y, et al. Polygenic prediction via Bayesian regression and continuous shrinkage priors. Nat Commun 2019;10:1776. doi:10.1038/s41467-019-09718-5

28 McHugh ML. Interrater reliability: The kappa statistic. Biochem Med (Zagreb) 2012;22:276–82. doi:10.11613/BM.2012.031

29 Hezel DM, Rose SV, Simpson HB. Delay to diagnosis in OCD. J Obsessive Compuls Relat Disord 2022;32:100709. doi:10.1016/j.jocrd.2022.100709

30 Perris F, Cipolla S, Catapano P, et al. Duration of Untreated Illness in Patients with Obsessive-Compulsive Disorder and Its Impact on Long-Term Outcome: A Systematic Review. J Pers Med 2023;13. doi:10.3390/jpm13101453

31 Ahn-Robbins D, Grootendorst-van Mil NH, Chang C-K, et al. Prevalence and Correlates of Obsessive-Compulsive Symptoms in Individuals With Schizophrenia, Schizoaffective Disorder, or Bipolar Disorder. J Clin Psychiatry 2022;83. doi:10.4088/JCP.21m14010

32 Kim J, Leonte KG, Chen ML, et al. Large language models outperform mental and medical health care professionals in identifying obsessive-compulsive disorder. npj Digital Med 2024;7:193. doi:10.1038/s41746-024-01181-x

33 All of Us Research Program Investigators, Denny JC, Rutter JL, et al. The “All of Us” Research Program. N Engl J Med 2019;381:668–76. doi:10.1056/NEJMsr1809937

