## Supplementary Material for "Development and validation of electronic health record-based ascertainment of obsessive-compulsive disorder cases and controls"

Figure S4. Quality control (QC) steps for the genotyping data of MGB Biobank individuals using PLINK. (a) Demonstration of QC steps for individuals genotyped on 9 batches on MEGA, MEGAEX, and MEG genotyping platforms; (b) Demonstration of QC steps for individuals genotyped on 4 batches on GSA genotyping platforms. . 26

### APPENDIX S1: DATA SOURCES

#### S1.1 Vanderbilt University Medical Center (VUMC)

VUMC is a large tertiary care and academic medical center located in Nashville, Tennessee, with a catchment area spanning from southern Kentucky to northern Alabama. Its clinical data were obtained from the Synthetic Derivative (SD), a de-identified electronic health record (EHR) data warehouse containing clinical information extracted from health records of over 2.2 million individuals in a searchable form covering the past 20 years. Available data types include (but are not limited to) structured data including ICD-9 and ICD-10 codes, PheWAS, CPT codes, administered medications, laboratory values, and vitals as well as unstructured data including clinical communications (including discharge summaries, H+P, progress notes, other communication between care providers, or between providers and patient, clinical reports including those from radiology, pathology, rehabilitation), problem lists, and family history. The SD interface allows users to search data extracted from most of the major health information databases at Vanderbilt including StarPanel and the enterprise data warehouse (EDW), which is a data warehouse integrating data from EPIC, Medipac, and Horizon Export Orders (WIZ). The search interface allows the user to input basic clinical and demographic information, such as ICD 9/10 codes, CPT procedure codes, medications, lab values, age, and gender, and returns de-identified data to the user for review and selection.

The clinical data were linked to BioVU (https://victr.vanderbilt.edu/pub/biovu), which is Vanderbilt University's repository of de-identified DNA extracted from discarded blood collected during routine clinical testing at VUMC [1]. It accrues 500-1000 samples per week, totalling more than 300,000 DNA samples as of January 2023. The combination of these two data resources allows for genome-phenome analysis at a scale that is among the most powerful in the United States. The VUMC Institutional Review Board oversees BioVU and approved this project (IRB201609).

##### S1.2 Mass General Brigham (MGB)

MGB is a large healthcare system providing clinical care to patients in Massachusetts, USA. Its clinical data were obtained from Research Patient Data Registry (RPDR) [2], which is a centralized data registry of clinical information from multiple EHR systems within MGB. The RPDR database covers more than 7 million patients with over 3 billion records from more than 8 hospitals, including two major teaching hospitals, namely Massachusetts General Hospital and Brigham and Women's Hospital, covering a wide range of patient characteristics including demographics, diagnoses, medications, procedures, lab tests and clinical notes. We queried the RPDR for all visits (i.e. inpatient, outpatient and emergency department visits) from occurring between Dec 1997 and January 2021 who met the following data floor: at least three visits since 2005, occurring more than 30 days apart, with at least one clinical note. The clinical notes used in this study included progress notes, discharge summaries, Epic ambulatory visit notes and Longitudinal Medical Record (LMR) visit notes. The MGB Biobank (MGBB) has recruited over 145,000 patients to date, providing samples and data that have supported more than 550 studies.

#### APPENDIX S2: ALGORITHM DEVELOPMENT

The algorithm development process followed the steps outlined below:

1. Working with domain experts (PGC investigators and VUMC psychiatry and neurology):
   a. Identify ICD-9 and -10 codes for exclusion (structured data).
   b. Identify ICD-9 and -10 codes for inclusion (structured data, for the ICD branch of the algorithm).
   c. Identify diagnostic keywords (for the NLP branch of the algorithm).
   d. Identify treatment-related keywords (for the NLP branch of the algorithm).
   e. Identify medications commonly used to treat OCD (used by MedEx, for the NLP branch of the algorithm).
2. Determine logical order of operations and document algorithm pipeline in pseudocode.
   a. Period of editing (commenting by domain experts) and operationalizing (commenting by "BioVU Programmers").
3. Apply the algorithm to individuals in SD.
4. Chart review round 1 - Evaluation of 25 subjects (including 5 subjects not identified by the algorithm as OCD cases) by clinically trained reviewers (EM, PM), with calibration performed between reviewers following the chart review.
   a. The SD interface allows reviewers to search data extracted from major health information databases at Vanderbilt and provides de-identified data for review.

    b. Reviewers reached a consensus on handling cases with multiple comorbidities. It was established that the presence of multiple comorbidities was acceptable, and as long as there was evidence of OCD, the comorbid conditions would not be treated as exclusions.

5. Error analysis:
    a. Identification of common false positives (e.g., mention of keyword in context of family member)
6. Refine the algorithm by, for example, adding additional exclusion and inclusion criteria, and re-apply it to SD.
7. Chart review round 2 - Evaluation of a new set of 25 subjects by the same clinically trained reviewers, with both reviewers independently reviewing all charts, to establish inter-rater reliability (IRR).
    a. Combining Review 1 and Review 2 resulted in a total of 50 charts reviewed by both EM and PM.
    b. IRR metrics:
        i. Raw agreement = 0.91
        ii. Cohen's Kappa = 0.72
        iii. Reviewer-Averaged Positive Predictive Value (PPV) = 0.81
8. Chart review final round - In the third round of evaluation, each reviewer (EM, PM) independently evaluated 25 unique charts, with adjudication by two psychiatrists (JB, HH), resulting in a total of 50 charts reviewed. These included 45 algorithm-determined OCD cases and 5 non-cases.

a. PPV = 0.64

9. Combined metrics (PPV) across three rounds of chart review per criteria:

    a. ICD or NLP present = 0.72 (70/97)

    b. ICD present (regardless of NLP) = 0.73 (48/66)

    c. NLP present (regardless of ICD) = 0.79 (64/81)

    d. ICD and NLP present = 0.84 (42/50)

    e. ICD without keyword present = 0.37 (6/16)

    f. NLP without ICD present = 0.71 (22/31)

#### APPENDIX S3: NLP IMPLEMENTATION

At Vanderbilt, we developed a web service version of SecTag [3], MedEx [4], NegEx [5], and KnowledgeMap Concept Identifier [6] using both Simple Object Access Protocol (SOAP) and Representational State Transfer (REST) protocols. This has been integrated with the Konstanz Information Miner (KNIME) interface [7], an open-source data analytics, reporting and integration platform that can be used for analysis as well as visualization of the data. SecTag identifies various sections and subsections within medical records, allowing text to be properly attributed to specific elements (e.g., family history or past medical history). The KnowledgeMap Concept Indexer (KMCI), the underlying NLP engine applied to the SD, extracts biomedical concepts from unstructured clinical text and maps them to Unified Medical Language System (UMLS) concepts. KMCI applies part-of-speech tagging to develop a shallow sentence parse and performs variant generation and normalization using the SPECIALIST Lexicon (https://lhncbc.nlm.nih.gov/LSG/Projects/lexicon/current/web/index.html) and related tools. KMCI leverages probabilistic information and concept co-occurrence data derived from PubMed to resolve ambiguous strings, such as mapping “OCD” to the UMLS concept C0028768 (“Obsessive Compulsive Disorder”) in psychiatry clinic notes or to C0029421 (“Osteochondritis Dessicans”) in documents discussing cartilage repair (a treatment for joint defects). KMCI has demonstrated favorable performance compared to MetaMap [8] and has been validated across various clinical and educational contexts.

The NLP system at MGB was implemented using medspaCy [9], a clinical NLP toolkit within the Python spaCy framework (https://spacy.io/). As depicted in Figure S1, the system comprises the following steps: 1) sentence segmentation using RuSH [10]; 2) keyword extraction; 3) attribute detection, including negation and experiencer detection using the ConText algorithm [11]; and 4) post-processing. Mentions of OCD or CBT-related keywords attributed to negation or a family member (experiencer) were excluded.

#### APPENDIX S4: CHART REVIEW INSTRUCTIONS

You will review subjects to determine that they meet criteria for an OCD or OCDS diagnosis based on instructions below.

Review Instructions:

1. Access the subject record by selecting the GRID in the Subject Detail Data panel.
2. Review all available information when determining whether to confirm inclusion/exclusion and level of evidence (See below), including relevant documents, ICD-9 and ICD-10 codes, and clinical notes.
3. For each patient, review the SD Discover and document your findings (the context around the keywords, multiple instances of ICD-9/10 codes) in the provided spreadsheet.

Algorithm requirement:

- Everyone in the collection will have at least 1 instance of ICD code for OCD or indicated OC spectrum disorder (ICD-9 300.3, ICD-10 F42.*).
- Or - At least one mention of OCD, obsessive-compulsive disorder, as well as medication or behavioral therapy comment.

For each subject you will determine true OCD diagnosis, utilizing a flag to note evidence level.

Inclusions strict criteria flag

- Has more than two relevant ICD codes on two or more separate days.
- Or has at least one ICD code and at least one mention of diagnosis or treatment in clinical notes, problem lists, clinical communication, discharge summaries (e.g. "Mrs. X has been diagnosed with OCD", or "OCD is managed through medication and CBT").

Inclusions broad criteria flag

- Has at least one ICD code and at least one mention of obsessive-compulsive behaviors or symptoms or traits.
- Or has more than three independent mentions of OCD, OC symptoms, OCD traits, or OC behaviors in clinical notes or problem lists without an ICD code.

Things to avoid:

- One single ICD code, no other evidence in the chart.
- One mention of OCD in the problem list with no additional supporting clinical information and no ICD code.
- Mention of no history of diagnosis(dx).
- Ruled out OCD (i.e., negating phrase).
- OCD of a family member without mention of OCD of the patient.
- Colloquial mention of OCD.

Flags:

High Evidence:

- Psychology clinic note
- Psychiatry clinic note
- Neurology clinic note
- Student mental health clinic note

Mid-Evidence:

- Internal medicine clinic note
- General med/peds clinic note

Low Evidence:

- Emergency department note
- Non-psych specialty clinic note (i.e., orthopedics)
- ICD
- Problem list

Exclusion criteria:

- Negating clauses: “no OCD”, “OCD negative”, “unremarkable for OCD”.

- Remove if mention of any immediate family member (i.e., wife, husband, brother, sister, son, daughter) in context and NO other mention of OCD or ICD code.
- Remove if mention of “osteochondritis” in context and NO other mention of OCD or ICD code.
- Remove if first mention > 55 years of age.
- Remove if only mention is prior to 2000.

<u>Clinical Definition of OCD</u>

The DSM-5 (*Diagnostic and Statistical Manual of Mental Disorders, Fifth Edition*) provides clinicians with official definitions of and criteria for diagnosing mental disorders and dysfunctions. Although not all experts agree on the definitions and criteria set forth in the DSM-5, it is considered the "gold standard" by most mental health professionals in the United States.

DSM-5 Diagnostic Criteria for Obsessive-Compulsive Disorder (300.3)

A. Presence of obsessions, compulsions, or both:

Obsessions are defined by (1) and (2):

1. Recurrent and persistent thoughts, urges, or impulses that are experienced, at some time during the disturbance, as intrusive and unwanted, and that in most individuals cause marked anxiety or distress.

2. The individual attempts to ignore or suppress such thoughts, urges, or images, or to neutralize them with some other thought or action (i.e., by performing a compulsion).

Compulsions are defined by (1) and (2):

1. Repetitive behaviors (e.g., hand washing, ordering, checking) or mental acts (e.g., praying, counting, repeating words silently) that the individual feels driven to perform in response to an obsession or according to rules that must be applied rigidly.

2. The behaviors or mental acts are aimed at preventing or reducing anxiety or distress, or preventing some dreaded event or situation; however, these behaviors or mental acts are not connected in a realistic way with what they are designed to neutralize or prevent, or are clearly excessive.

*Note:* Young children may not be able to articulate the aims of these behaviors or mental acts.

B. The obsessions or compulsions are time-consuming (e.g., take more than 1 hour per day) or cause clinically significant distress or impairment in social, occupational, or other important areas of functioning.

C. The obsessive-compulsive symptoms are not attributable to the physiological effects of a substance (e.g., a drug of abuse, a medication) or another medical condition.

D. The disturbance is not better explained by the symptoms of another mental disorder (e.g., excessive worries, as in generalized anxiety disorder; preoccupation with appearance, as in body dysmorphic disorder; difficulty discarding or parting with possessions, as in hoarding disorder; hair pulling, as in trichotillomania [hair-pulling disorder]; skin picking, as in excoriation [skin-picking] disorder; stereotypies, as in stereotypic movement disorder; ritualized eating behavior, as in eating disorders; preoccupation with substances or gambling, as in substance-related and addictive disorders; preoccupation with having an illness, as in illness anxiety disorder; sexual urges or fantasies, as in paraphilic disorders; impulses, as in disruptive, impulse-control, and conduct disorders; guilty ruminations, as in major depressive disorder; thought insertion or delusional preoccupations, as in schizophrenia spectrum and other psychotic disorders; or repetitive patterns of behavior, as in autism spectrum disorder).

#### APPENDIX S5: QUALITY CONTROL IN GENETIC VALIDATION

##### S5.1 Vanderbilt University Medical Center (VUMC)

We obtained genotype information on 94,474 BioVU individuals genotyped on the Illumina MEGA EX array. Using PLINK v1.95 [12], genotypes were filtered for SNP and individual call rates, sex discrepancies, and excessive heterozygosity. We confirmed the absence of genotyping batch effects through logistic regression with batch as the phenotype. Autosomes were imputed to the HRC panel using Michigan Imputation Server 4 in five batches. After imputation, genotypes were converted to hard calls with PLINK using the default threshold settings. SNPs with multiple alleles or imputation quality less than $R^2$ of 0.3 were excluded. Next, SNPs with minor allele frequency less than 0.005 or genotyping rates less than 0.98 were excluded. Individuals with call rates less than 0.98 were excluded.

We ran a series of principal component analyses (PCA) to determine BioVU individuals with the greatest genetic similarity. First, we performed PCA using FlashPCA on BioVU combined with CEU, YRI, and CHB reference sets from 1000 Genomes Project Phase 3 (https://www.internationalgenome.org/category/phase-3/). Principal components were scaled so that the axes could be interpreted as proportions of genetic similarity to the reference populations. We selected BioVU individuals who were within 40% of the CEU cluster along the CEU-CHB axis and within 30% of the CEU cluster on the CEU-YRI axis, generating a once-PCA filtered 1KG-CEU-clustered dataset.

We then filtered the previously identified BioVU European cluster to identify individuals falling within the CEU, TSI, and GIH 1000 genomes populations, producing a twice filtered

1KG-EU-clustered dataset. Using the twice-filtered European set we conducted a series of SNP checks. First, we filtered individuals with IBS greater than 0.2 and calculated principal components to use as covariates. Next, we checked for imputation batch effects by conducting pairwise logistic regression of the five imputation batches using sex and top 10 principal components as covariates. SNPs with p-values less than 0.001 in the additive model were flagged. We then compared MAF between BioVU and the CEU reference population. Any SNPs with a MAF difference greater than 0.1 were flagged. SNPs with a Hardy-Weinberg Equilibrium p-value less than 10E-10 were flagged. Finally, the flagged SNPs from the batch effect, MAF difference, and HWE were excluded from the once-PCA filtered BioVU European set, resulting in 9,386,383 SNPs for analysis. The final dataset for genetic validation consisted of 676 (ICD or NLP) cases and 46,677 controls.

**S5.2 Mass General Brigham (MGB)**

A total of 53,853 MGB Biobank individuals were genotyped in 13 batches using four different Illumina genotyping array platforms: Multi-Ethnic Genotyping Array (MEGA, batch 1), Expanded Multi-Ethnic Genotyping Array (MEGAEX, batch 2), Multi-Ethnic Global (MEG, batch 3-9), or Global Screening Array (GSA, batch 10-13). Samples from batches 8 and 9 were excluded due to the small number of cases (15 and 10 OCD cases identified by ICD-or-NLP, respectively). Within each batch, standard quality control was conducted on individuals and SNP genotyping data using PLINK v1.95. Individuals with call rates < 98%, ambiguous genomic sex, sex discrepancy between ascertainment record and genomic sex, inbreeding coefficient |F|>0.2, or related samples with PI_HAT > 0.2 were removed. SNP quality control included the exclusion of SNPs with genotyping rate < 98%, differential missingness between cases and controls > 0.02, minor allele frequency (MAF) < 0.01, strand-ambiguous SNPs, and SNPs with $P < 1x10^{-6}$ in Hardy Weinberg Equilibrium

(HWE) tests among controls. PCA was performed to identify individuals of European ancestry and non-European individuals were removed.

After the quality control steps within each batch, individuals genotyped on MEGA, MEGAEX, and MEG platforms were merged into the MEG set, retaining only SNPs that passed quality control in all 7 batches. Pairwise batch effect tests (21 tests in total) were performed using genome-wide logistic regression, treating controls from one batch as cases and controls from another batch as controls. PCA components were included in the logistic regression to control for population stratification, and SNPs with $P<1x10^{-5}$ for batch effects were excluded. Similarly, individuals genotyped in the 4 GSA batches were merged into the GSA set, and SNPs exhibiting batch effects were removed using the same approach. Identity By Descent (IBD) analysis was performed on all individuals, and duplicates and relatives across the two datasets were excluded.

After quality control, controls were matched to cases at a 4:1 ratio based on gender, genotyping batch, age (within 10 years), and genetic proximity determined by the first two PCA components. The genotyped data of cases and matching controls were phased using SHAPEIT2 [13] and imputed in Minimac3 [14] using the HRC reference panel (release 1.1). The MEG and GSA datasets were phased and imputed separately. Post-imputation, SNPs with MAF>0.01 and INFO scores between 0.8 and 1.2 were retained for OCD polygenic risk score (PRS) calculation.

#### SUPPLEMENTARY TABLES

Table S1: Number of controls and cases by array platforms and batches at MGB.

Table S2. Demographic composition of the resulting OCD cohorts.

Table S1. Number of controls and cases by array platforms and batches at MGB.

| Batch | Array | Controls | Cases |
|---|---|---|---|
| 1 | MEGA | 4554 | 97 |
| 2 | MEGAEX | 4948 | 105 |
| 3 | MEG | 4401 | 111 |
| 4 | MEG | 4538 | 143 |
| 5 | MEG | 4914 | 168 |
| 6 | MEG | 4606 | 131 |
| 7 | MEG | 4337 | 109 |
| 8 | MEG | 817 | 15 |
| 9 | MEG | 133 | 10 |
| 10 | GSA | 10744 | 393 |
| 11 | GSA | 6539 | 152 |
| 12 | GSA | 5028 | 93 |
| 13 | GSA | 19447 | 475 |

Table S2. Demographic composition of the resulting OCD cohorts.

| **Setting** | **ICD-or-NLP (VUMC)** | **ICD-and-NLP (VUMC)** | **ICD-or-NLP (MGB)** | **ICD-and-NLP (MGB)** |
|---|---|---|---|---|
| Genotyped *N* | 1057 | 515 | 1379 | 335 |
| Age, mean (SD) | 45.34 (16.83) | 45.27 (17.79) | 50.46 (15.71) | 44.77 (10.62) |
| Gender: *N* (%) | | | | |
| *Female* | 631 (59.70) | 285 (55.34) | 882 (63.96) | 214 (63.88) |
| *Male* | 426 (40.30) | 230 (44.66) | 497 (36.04) | 121 (36.12) |
| Self-reported Race: *N* (%) | | | | |
| *Asian* | 13 (1.23) | 6 (1.17) | 27 (1.96) | 3 (0.90) |
| *Black/African American* | 57 (5.39) | 25 (4.85) | 62 (4.50) | 7 (2.09) |
| *White* | 965 (91.30) | 476 (92.43) | 1187 (86.08) | 304 (90.75) |
| *Other* | 4 (0.38) | 3 (0.58) | 84 (6.09) | 15 (4.48) |
| *Unknown* | 18 (1.70) | 5 (0.97) | 19 (1.38) | 6 (1.79) |
| Ethnicity: *N* (%) | | | | |

| | | | | |
|---|---|---|---|---|
| *Hispanic* | 23 (2.18) | 9 (1.75) | 32 (2.32) | 6 (1.79) |
| *Non-Hispanic* | 1034 (97.82) | 506 (98.25) | 1347 (97.68) | 329 (98.21) |
| Public payer: *N* (%) | | | | |
| *Yes* | 543 (51.37) | 276 (53.59) | 806 (58.45) | 193 (57.61) |
| *No* | 514 (48.63) | 239 (46.41) | 573 (41.55) | 142 (42.39) |
| Hospital usage: Median (IQR) | | | | |
| *EHR length (in years)* | 8.30 (12.07) | 8.33 (12.16) | 16.43 (12.45) | 16.34 (11.72) |
| *Visit count* | 70 (119) | 78 (129) | 315.0 (427.5) | 330.0 (411.5) |
| *ICD count* | 188 (385) | 213 (408) | 686.0 (1101.5) | 669.0 (1033.0) |
| *CPT count* | 191 (325) | 219 (328) | 502.0 (750.5) | 489.0 (758.5) |
| *RxNorm count* | 110 (148.25) | 120 (156.5) | 481.0 (824.0) | 539.0 (842.0) |
| *LOINC count* | 153 (127.5) | 159 (127) | 493.5 (496.25) | 497.0 (545.5) |
| *Note count* | 1117.5 (2220.75) | 1187 (2239.5) | 321.0 (492.0) | 335.0 (502.0) |

**SUPPLEMENTARY FIGURES**

Figure S1: The steps for processing notes and classifying patient-level OCD status at MGB.

Figure S2: Data entry interface used for chart review at MGB determining the OCD status of a given patient.

Figure S3: Venn diagrams summarizing the OCD case counts, identified by the algorithm and genotyped in (a) BioVU at VUMC and (b) the MGB Biobank at MGB.

Figure S4: Quality control (QC) steps for the genotyping data of MGB Biobank individuals using PLINK. (a) Demonstration of QC steps for individuals genotyped on 9 batches on MEGA, MEGAEX, and MEG genotyping platforms; (b) Demonstration of QC steps for individuals genotyped on 4 batches on GSA genotyping platforms.

Figure S1. The steps for processing notes and classifying patient-level OCD status at MGB.

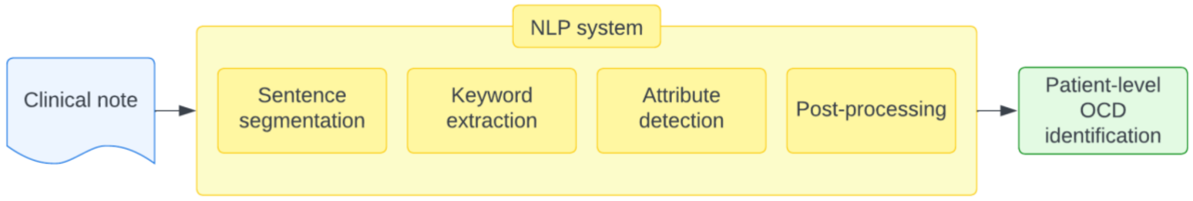

Figure S2. Data entry interface used for chart review at MGB determining the OCD status of a given patient.

Record ID ______________________________

**Patient Information**

Patient MRN ______________________________

Patient Date of Birth (if available) ______________________________

Patient Gender (if available)

- ○ Female
- ○ Male
- ○ Not reported

**Chart Review:**

Referring to the information documented in the chart, please classify the patient as one of the options below.

You should classify this patient as a case of OCD only if you believe that it is more likely than not that this patient has OCD based on the documented history available.

Evidence of diagnosis could include:

Mention of diagnosis or treatment in clinical notes, clinical communication, discharge summaries (e.g. □Mrs. X has been diagnosed with OCD□, or □OCD is managed through medication and CBT□). OCD medication (eg. high dose SSRI ( clomipramine), in the absence of a depressive or anxiety disorder or PTSD) Mention of obsessive-compulsive behaviors or symptoms or traits >2 independent mentions of OCD, OC symptoms, OCD traits, or OC behaviors in clinical notes or problem list without an ICD code. Note: Only 1 mention in the context of Past Medical History or the opening line of an HPI entry (similar to PMH) or in Problem List may be counted. At least 1 mention must come from a source other than these.

- ○ OCD
- ○ Not OCD

Please document the highest level of evidence present. For example, if there is evidence from an emergency room note and a psychiatry clinic note, this would be considered a □high evidence□ case and would receive the corresponding □high evidence□ flag.

High Evidence:

Psychology clinic note Psychiatry clinic note (outpatient or inpatient) Neurology clinic note (outpatient or inpatient) Student mental health clinic note Developmental medicine Mid-Evidence:

Internal medicine clinic note General med/peds clinic note Inpatient adult or peds General Medicine notes Low Evidence:

Emergency department note Non-psych specialty clinic note (i.e., orthopedics) Problem list

- ○ High Evidence
- ○ Mid Evidence
- ○ Low Evidence

Does this patient have a PANDAS diagnosis?

- ○ Yes
- ○ No

Please provide a 1-3 sentence summary of your determination.

NOTE: No protected health information (PHI) should be recorded in this form.

Figure S3. Venn diagrams summarizing the OCD case counts, identified by the algorithm and genotyped in (a) BioVU at VUMC and (b) the MGB Biobank at MGB.

(a) Summary of BioVU OCD case counts

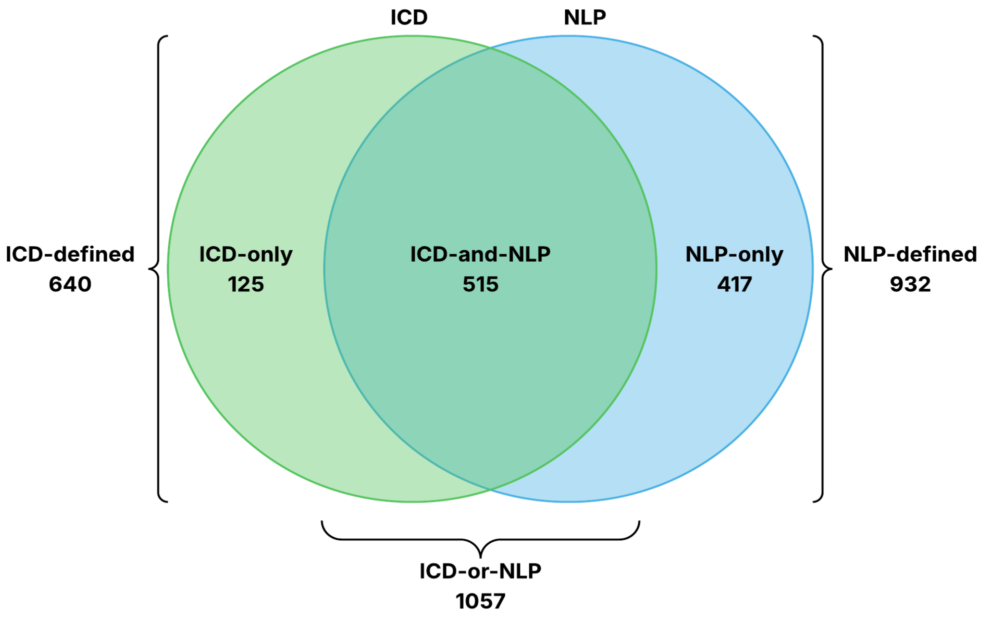

(b) Summary of MGBB OCD case counts

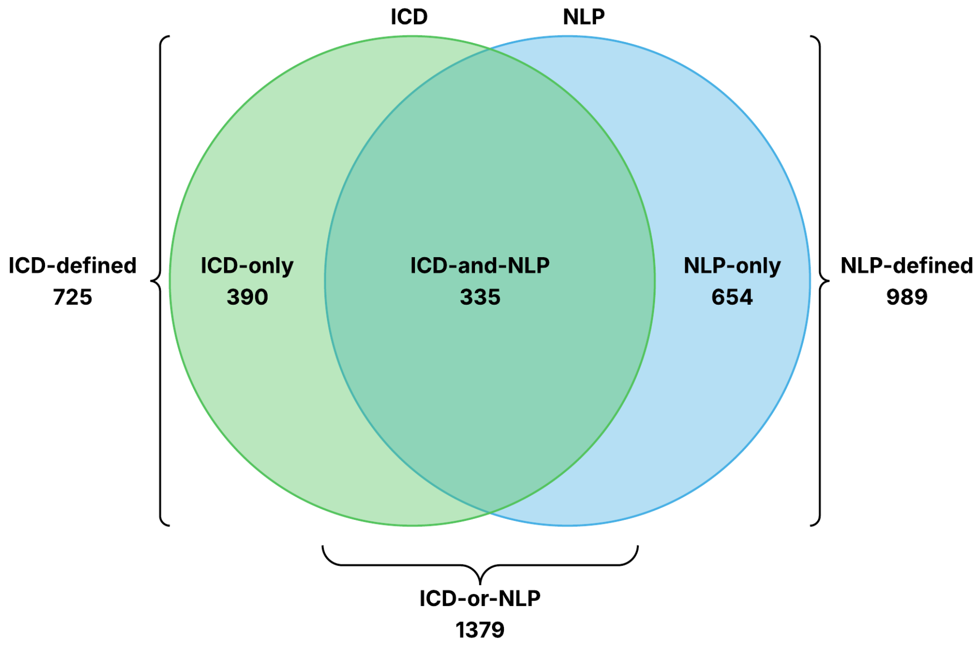

Figure S4. Quality control (QC) steps for the genotyping data of MGB Biobank individuals using PLINK. (a) Demonstration of QC steps for individuals genotyped on 9 batches on MEGA, MEGAEX, and MEG genotyping platforms; (b) Demonstration of QC steps for individuals genotyped on 4 batches on GSA genotyping platforms.

HWE: Hardy-Weinberg equilibrium test; MAF: minor allele frequency; HET |F|: inbreeding coefficient (F) measuring the level of heterozygosity for an individual.

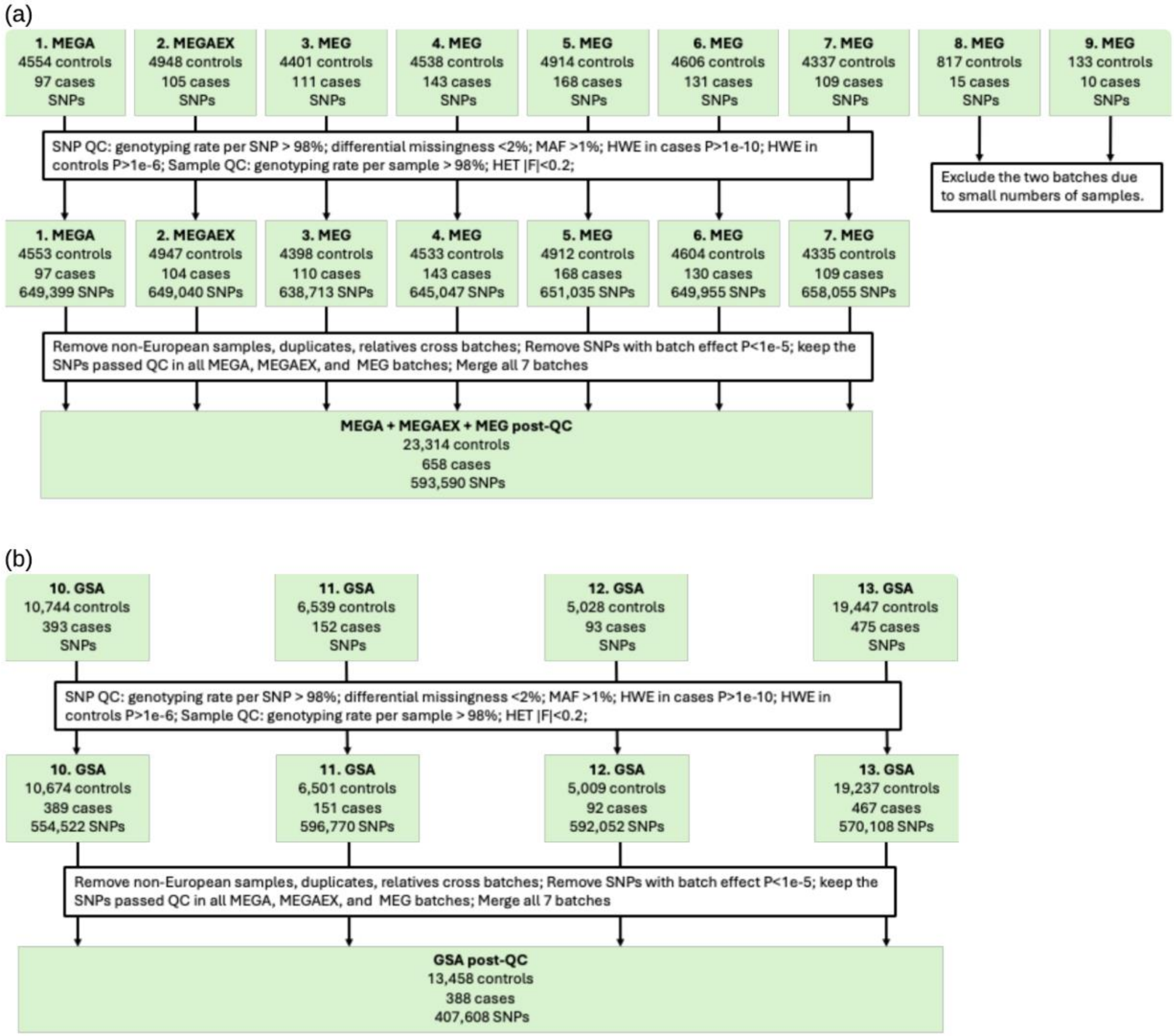

#### REFERENCES

1 Roden DM, Pulley JM, Basford MA, *et al.* Development of a large-scale de-identified DNA biobank to enable personalized medicine. *Clin Pharmacol Ther* 2008;**84**:362–9. doi:10.1038/clpt.2008.89

2 Nalichowski R, Keogh D, Chueh HC, *et al.* Calculating the benefits of a Research Patient Data Repository. *AMIA Annu Symp Proc* 2006;:1044.

3 Denny JC, Spickard A, Johnson KB, *et al.* Evaluation of a method to identify and categorize section headers in clinical documents. *J Am Med Inform Assoc* 2009;**16**:806–15. doi:10.1197/jamia.M3037

4 Xu H, Stenner SP, Doan S, *et al.* MedEx: a medication information extraction system for clinical narratives. *J Am Med Inform Assoc* 2010;**17**:19–24. doi:10.1197/jamia.M3378

5 Chapman WW, Bridewell W, Hanbury P, *et al.* A simple algorithm for identifying negated findings and diseases in discharge summaries. *J Biomed Inform* 2001;**34**:301–10. doi:10.1006/jbin.2001.1029

6 Denny JC, Irani PR, Wehbe FH, *et al.* The KnowledgeMap project: development of a concept-based medical school curriculum database. *AMIA Annu Symp Proc* 2003;:195–9.

7 Berthold MR, Cebron N, Dill F, *et al.* KNIME: The Konstanz Information Miner. In: Preisach C, Burkhardt H, Schmidt-Thieme L, *et al.*, eds. *Data Analysis, Machine Learning and Applications*. Berlin, Heidelberg: : Springer Berlin Heidelberg 2008. 319–26. doi:10.1007/978-3-540-78246-9_38

8 Aronson AR, Lang F-M. An overview of MetaMap: historical perspective and recent advances. *J Am Med Inform Assoc* 2010;**17**:229–36. doi:10.1136/jamia.2009.002733

9 Eyre H, Chapman AB, Peterson KS, *et al.* Launching into clinical space with medspaCy: a new clinical text processing toolkit in Python. *AMIA Annu Symp Proc* 2021;**2021**:438–47. doi:10.48550/arxiv.2106.07799

10 Shi J, Mowery D, Doing-Harris KM, *et al.* RuSH: a Rule-based Segmentation Tool Using Hashing for Extremely Accurate Sentence Segmentation of Clinical Text. *American Medical Informatics Association Annual Symposium* 2016.

11 Chapman WW, Chu D, Dowling JN. ConText: An algorithm for identifying contextual features from clinical text. In: *Proceedings of the Workshop on BioNLP 2007 Biological, Translational, and Clinical Language Processing - BioNLP '07*. Morristown, NJ, USA: : Association for Computational Linguistics 2007. 81. doi:10.3115/1572392.1572408

12 Purcell S, Neale B, Todd-Brown K, *et al.* PLINK: a tool set for whole-genome association and population-based linkage analyses. *Am J Hum Genet* 2007;**81**:559–75. doi:10.1086/519795

13 Delaneau O, Marchini J, Zagury J-F. A linear complexity phasing method for thousands of genomes. *Nat Methods* 2011;**9**:179–81. doi:10.1038/nmeth.1785

14 Li Y, Willer C, Sanna S, *et al.* Genotype imputation. *Annu Rev Genomics Hum Genet* 2009;**10**:387–406. doi:10.1146/annurev.genom.9.081307.164242
